# Omicron B.1.1.529 variant infections associated with severe disease are uncommon in a COVID-19 under-vaccinated, high SARS-CoV-2 seroprevalence population in Malawi

**DOI:** 10.1101/2022.08.22.22279060

**Authors:** Upendo L. Mseka, Jonathan Mandolo, Kenneth Nyoni, Oscar Divala, Dzinkambani Kambalame, Daniel Mapemba, Moses Kamzati, Innocent Chibwe, Marc Y. R. Henrion, Kingsley Manda, Deus Thindwa, Memory Mvula, Bright Odala, Raphael Kamng’ona, Nelson Dzinza, Khuzwayo C. Jere, Nicholas Feasey, Antonia Ho, Abena S. Amoah, Melita Gordon, Todd D Swarthout, Amelia Crampin, Robert S. Heyderman, Matthew Kagoli, Evelyn Chitsa-Banda, Collins Mitambo, John Phuka, Benson Chilima, Watipaso Kasambara, Kondwani C. Jambo, Annie Chauma-Mwale

## Abstract

**Background:** The B.1.1.529 (Omicron) variant of severe acute respiratory syndrome coronavirus 2 (SARS-CoV-2) has resulted in the fourth COVID-19 pandemic wave across the southern African region, including Malawi. The seroprevalence of SARS-CoV-2 antibodies and their association with epidemiological trends of hospitalisations and deaths are needed to aid locally relevant public health policy decisions.

**Methods:** We conducted a population-based serosurvey from December 27, 2021 to January 17, 2022, in 7 districts across Malawi to determine the seroprevalence of SARS-CoV-2 antibodies. Primary sampling units (PSU) were selected using probability proportionate to the number of households based on the 2018 national census, followed by second-stage sampling units that were selected from listed households. A random systematic sample of households was selected from each PSU within the 7 districts. Serum samples were tested for antibodies against SARS-CoV-2 receptor binding domain using WANTAI SARS-CoV-2 Receptor Binding Domain total antibody commercial enzyme-linked immunosorbent assay (ELISA). We also evaluated COVID-19 epidemiologic trends in Malawi, including cases, hospitalizations and deaths from April 1, 2021 through April 30, 2022, collected using the routine national COVID-19 reporting system.

**Results:** Serum samples were analysed from 4619 participants (57% female; 65% aged 14 to 50 years), of whom 1018 (22%) had received a COVID-19 vaccine. The overall assay-adjusted seroprevalence was 86.3% (95% confidence interval (CI), 85.1% to 87.5%). Seroprevalence was lowest among children <13 years of age (66%) and highest among adults 18 to 50 years of age (82%). Seroprevalence was higher among vaccinated compared to unvaccinated participants (96% vs. 77%; risk ratio, 6.65; 95% CI, 4.16 to 11.40). Urban residents were more likely to test seropositive than those living in rural settings (91% vs. 78%; risk ratio, 2.81; 95% CI, 2.20 to 3.62). National COVID-19 data showed that at least a two-fold reduction in the proportion of hospitalisations and deaths among the reported cases in the fourth wave compared to the third wave (hospitalization, 10.7% (95% CI, 10.2 to 11.3) vs 4.86% (95% CI, 4.52 to 5.23), p<0.0001; deaths, 3.48% (95% CI, 3.18 to 3.81) vs 1.15% (95% CI, 1.00 to 1.34), p<0.0001).

**Conclusion:** We report reduction in proportion of hospitalisations and deaths from SARS-CoV-2 infections during the Omicron variant dominated wave in Malawi, in the context of high SARS-CoV-2 seroprevalence but low COVID-19 vaccination coverage. These findings suggest that COVID-19 vaccination policy in high seroprevalence settings may need to be amended from mass campaigns to targeted vaccination of at-risk populations.

## Introduction

The B.1.1.529 (Omicron) variant of severe acute respiratory syndrome coronavirus 2 (SARS-CoV-2) was first reported in South Africa on 25^th^ November 2021. Within approximately 25 days the Omicron variant was responsible for over 90% of the new clinical cases in South Africa, compared to 80% with the Delta variant 100 days after its emergence in the same setting ^1^. Compared to wild type SARS-CoV-2, the Omicron infection rate was four times higher and the Delta rate was twice as high ^2^. Genomic sequencing of the Omicron variant has revealed a wide range of non-synonymous mutations ^3,4^, with 30 mutations identified within its spike protein that have been linked to its high transmission rate and greater immune evasion of neutralizing antibodies ^5-7^. Despite the Omicron variant being associated with higher transmissibility, coronavirus disease (COVID-19) cases caused by this variant have mostly been less severe than previous variants in both low middle income and high-income settings ^1,8^.

The COVID-19 pandemic trajectory in Malawi has mirrored that of South Africa ^8^. Post emergence of variants of concern in South Africa, Malawi has experienced three COVID-19 waves, caused by Beta, Delta and Omicron ^8^. As of July 2022, the number of reported COVID-19 clinical cases and deaths in Malawi were 86,823 and 2,651, respectively ^9^. However, in the context of a low COVID-19 vaccination coverage (4.5%) in Malawi ^9^, data from a serosurvey among blood donors showed very high seropositivity (>65%) by July 2021 ^10^, suggesting widespread community transmission and a huge under-ascertainment of SARS-CoV-2 infections and possibly deaths. A recent sero-epidemiological study in South Africa found high SARS-CoV-2 seropositivity in COVID-19 unvaccinated (68%) and vaccinated (93%) individuals prior to the onset of the Omicron wave, and this was associated with reduced incidence of COVID-19 hospitalisation, recorded deaths and excess deaths thereafter ^1^. Consequentially, South Africa has taken a more pragmatic approach to COVID-19, accepting the limited success in preventing infections and allowing for policy development that is built on minimising severe COVID-19, while balancing the direct and indirect societal effects of COVID-19 ^11,12^. However, South Africa is the richest economy on the continent ^13^, had stringent lockdowns and has epidemiological profiles of communicable and non-communicable co-morbidities that differ from elsewhere in the region ^11,12,14^. To inform public health policy decisions in very low COVID-19 vaccine coverage and resource-constrained countries such as Malawi, further data are urgently needed.

We, therefore, undertook a population-based serosurvey in Malawi that was conducted from December 27, 2022 to January 17, 2022, during the Omicron-dominant fourth wave. Furthermore, we link this data to the national COVID-19 epidemiologic trends in Malawi, including clinical cases, hospitalizations and deaths, from April 1, 2021 through April 30, 2022.

## Methods

### Study design

The study used a stratified multistage probability sample design, with strata defined by five health zones, primary sampling units (PSU) defined by enumeration areas (EAs) within strata, second-stage sampling units defined by households within EAs, and finally eligible persons within households.

Following a sample size calculation, the intention was to sample 12 of the 29 districts of Malawi but due to logistical and operational challenges, only 7 of the 12 districts, namely Kasungu, Dowa, Dedza, Lilongwe, Zomba, Machinga, and Blantyre, were included in the study participant recruitment phase from 27^th^ December 2021 to 17^th^ January 2022. The 7 districts harbor 45% of the population of Malawi (7,911,594 of 17,563,749 persons) ^15^. The other 5 districts that constitute 13% of the Malawi’s population (2,221,932 of 17,563,749 persons) ^15^ were included in an earlier serosurvey. The districts of Lilongwe (2,626,901), Zomba (851,737) and Blantyre (1,251,484) have both rural and urban areas, and harbor 3 of Malawi’s main 4 cities.

### Sample size calculation

We assumed 5% prevalence of SARS-CoV-2 antibodies, a margin of error of ±5 %, 95% precision of estimates, and a design effect of 1.26. Using the following formula, we calculated a sample size (n) of 7,744 persons.

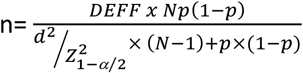

Where;

Population size (N): 17,563,749

Hypothesized estimate of the prevalence of SARS-CoV-2 (*p*): 5%±-5

Confidence limits as % of 100(absolute +/- %) (d): 5%

Design effect (DEFF): 1.26

Malawi has five health zones in three administrative regions (North, Southeast, Southwest, Central East, and Central West). Districts were selected across Malawi using simple randomisation within the health zones, choosing 2 districts per health zone, making a total of 10 districts, namely Nkhata Bay, Mzimba North, Kasungu, Dowa, Ntcheu, Dedza, Zomba, Machinga, Nsanje, and Neno. Additionally, the two largest cities of Blantyre and Lilongwe were added, totaling 12 districts. The PSUs were selected with probabilities proportionate to the number of households in the EA based on the 2018 census ^15^. The second-stage sampling units were selected from lists of households complied by trained staff for each of the sampled PSUs. Upon completion of the listing process, a random systematic sample of households was selected from each PSU within each health zone to the extent feasible. Within the sampled households, all eligible participants were included in the study sample for data collection. Assuming an average household size of 4.4 from the national census ^15^, the estimated households to be recruited was 1,760. However, for the 7 districts, the expected number of households to be recruited was 1,420 and a total sample size of 6,248 persons.

Inclusion criteria included being aged ≥5 years, residing in the sampled household, providing informed written consent (or consent from parent/guardian and assent if aged 12 to 17 years), willing to provide the required samples, and having no recent history of nose bleeding. After the consenting process, a standard COVID-19 screening tool was implemented using electronic data capturing (Open Data Kit, ODK), including demographic data, socio-economic status, comorbidities, COVID-19 symptoms, and COVID-19 vaccination status from each enrolled individual. COVID-19 vaccination status was ascertained from all participants using the individual national COVID-19 vaccination card.

The National Health Sciences Research Committee (NHSRC) (Protocol #21/02/2671) gave ethical approval for this work and further authorization was also obtained from the District Health Offices of the participating districts. Community sensitization within each district was implemented through community leadership and study staff prior to data collection. All participants provided written informed consent including assent for children; those who were approached were informed that they were free to decline participation or withdraw at any time without any negative repercussion.

### Sample collection and processing

Peripheral venous blood samples (4ml) were obtained from all enrolled participants. Samples were transported to the district laboratory, where serum was separated by density centrifugation, aliquoted and kept at 2-8°C for transportation to the main storage facility at the National Public Health Reference Laboratory (NPHRL) repository. Serum was archived at −80° C.

### Serological analysis

SARS-CoV-2 Receptor Binding Domain protein immunoglobulin were measured qualitatively in sera using the WANTAI SARS-CoV-2 total antibody commercial ELISA kit as described previously ^10^, following the manufacturer’s instructions (Beijing Wantai Biological Pharmacy Enterprise Co., Ltd., China; WS-1096). The sensitivity and specificity of the assay as independently validated are 94% [95% *CI* 0.91 to 0.97]) and 100% [95% *CI* 0.99 to 1,00]), respectively ^16^. The results are expressed as a ratio which is calculated by dividing the optical densities of the test sample by those of assay cutoff (mean OD of three internal negative calibrators + 0.16). Specimens giving a ratio of < 0.9 were reported as negative for this assay, a ratio of > 1.1 were reported as positive, and a ratio between 0.9 and 1.1 were reported as borderline. Samples with borderline results were retested using a confirmatory assay described below.

### National COVID-19 data sources

Data regarding daily cases, hospitalizations, and deaths were sourced from the Public Health Institute of Malawi (PHIM), from 1^st^ April 2021 through 30^th^ April 2022 ^9^. In brief, the routine national COVID-19 reporting system works as follows: data from multiple sources at district level covering a 24-hour period (6am-6am) are sent to national level through the national Public Health Emergency Operations Centre (PHEOC). The data undergoes a triple verification system against three sources, surveillance, laboratory, and case management, and then released to the public.

Cases included asymptomatic and symptomatic infections with SARS-CoV-2 confirmed by either a nucleic acid amplification assay or a rapid antigen test. Hospitalizations included admissions for SARS-CoV-2 infection, as well as admissions for other illnesses in which SARS-CoV-2 infection was incidentally identified on routine screening at the time of admission. Death attributable to COVID-19 was defined as a death resulting from a clinically compatible illness in a probable or confirmed COVID-19 case unless there is a clear alternative cause of death that was not related to COVID-19 disease (e.g., trauma), according to Malawi national guidelines.

### Statistical methods

Statistical analyses were performed using R statistical package (version 4.1.0) ^17^. Categorical variables were summarized as frequencies and percentages. Seropositivity was calculated as the proportion of those who tested positive among all those tested. The overall seroprevalence of the SARS-CoV-2 antibodies was adjusted for both the assay’s sensitivity (94% [95% *CI* 0.91 to 0.97]) and specificity (100% [95% *CI* 0.99 to 1,00]) ^16^. The χ^2^ test or Fisher’s exact test was used to compute the proportions of participants by number of COVID-19 vaccine doses received across several variables. A multivariate logistic regression model was developed to investigate the factors associated with SARS-CoV-2 seropositivity. The factors included age, sex, occupational exposure and COVID-19 vaccination, which were chosen based on known predictors of seropositivity. A p-value of <0.05 was considered statistically significant.

## Results

### Participant demographic and household characteristics

We enrolled 4,639 participants from 1,415 households, with 4,619 samples from the participants analysed (**Figure 1**). This constituted 74% of the target sample of 6,248 persons. The response rate was generally high across all districts however reasons for refusal included cultural and religious beliefs against drawing of blood, mistaking the serosurvey for COVID-19 vaccination programmes, lack of incentive and general fear towards COVID-19 related activities. Demographic and household characteristics, comorbidities, participant-reported HIV status, and COVID-19 vaccination status are shown in **Table 1**. In brief, 60% (2,751/4,619) of the participants were between the ages of 18 to 50 years, 57% (2,617/4,619) were female, and 7.8% (361/4619) reportedly had had a chronic illness other than HIV infection.

**Table 1.** Participant demographics and clinical history.

| Characteristic | N = 4,619<sup>†</sup> |
| --- | --- |
| **Sex** |  |
| Female | 2,617 (57%) |
| Male | 2,002 (43%) |
| **Age group** |  |
| <13 yrs | 187 (4.4%) |
| 13 to 17 yrs | 230 (5.5%) |
| 18 to 50 yrs | 2,751 (65%) |
| >50 yrs | 1,043 (25%) |
| Unknown | 408 |
| **COVID-19 Vaccination status** |  |
| 0 doses | 3,606 (78%) |
| 1 dose | 521 (11%) |
| 2 doses | 492 (11%) |
| **SARS-CoV-2 serostatus** |  |
| Negative | 865 (19%) |
| Positive | 3,754 (81%) |
| **Symptoms in the last 2 weeks** |  |
| Yes | 1,747 (38%) |
| No | 2,870 (62%) |
| Unknown | 2 (<0.1%) |
| **Occupation** |  |
| Irregular | 522 (11%) |
| Regular | 364 (7.9%) |
| Unwaged | 3,733 (81%) |
| **Education level** |  |
| Primary | 2,086 (45%) |
| Secondary | 1,038 (22%) |
| Tertiary | 232 (5.0%) |
| No formal education | 1,263 (27%) |
| **Comorbidities** |  |
| Yes | 361 (7.8%) |
| No | 4,258 (92%) |
| **HIV status** |  |
| HIV- | 4,457 (96%) |
| HIV+ | 162 (3.5%) |
| **Economic zone** |  |
| Urban | 1,373 (30%) |
| Rural | 1,156 (25%) |
| *Other | 2,090 (45%) |
| **Location** |  |
| Blantyre | 768 (17%) |
| Dedza | 527 (11%) |
| Dowa | 441 (9.5%) |
| Kasungu | 645 (14%) |
| Lilongwe | 1,327 (29%) |
| Machinga | 477 (10%) |
| Zomba | 434 (9.4%) |
| <sup>†</sup> n (%) |  |

\* includes other districts apart from city-harboring districts of Lilongwe, Blantyre and Zomba

**Figure 1.**
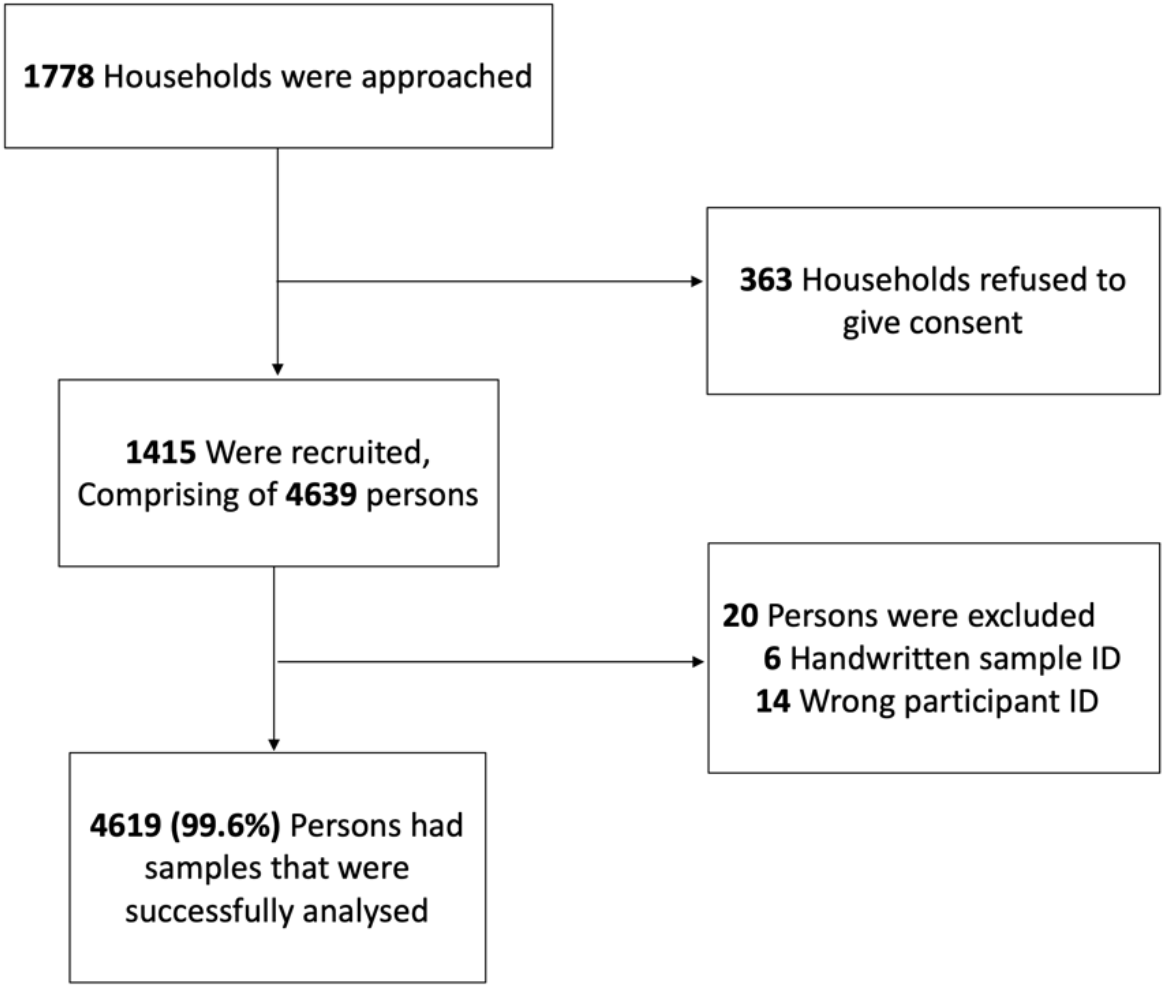
Survey participant recruitment. The survey was conducted between 27^th^ December 2021 to 17^th^ January 2022. The survey included primary sampling units (PSU), which were selected with probabilities proportionate to the number of households based on the 2018 census. The second-stage sampling units were selected from lists of households sampled PSUs, then a random systematic sample of households was selected from each PSU within each health zone to the extent feasible.

### SARS-CoV-2 seroprevalence estimates

Among all participants, the overall assay sensitivity and specificity adjusted seroprevalence was 86.3% (95% confidence interval (CI), 85.1% to 87.5%). The adjusted seroprevalence was heterogeneous across the districts, ranging from 80.4% (95% CI, 75.9% to 84.6%) in Dowa, to 93.7% (95% CI, 91.1% to 96.0%) in Blantyre (**Table 2**).

**Table 2.** SARS-CoV-2 antibody seroprevalence estimates.

|  | *n* | *Seroprevalence* | *95% Confidence Interval* | *Seroprevalence \*(adjusted)* | *95% Confidence Interval \*(adjusted)* |
| --- | --- | --- | --- | --- | --- |
| *All* | 3755/4619 | 81.3% | 80.1% to 82.4% | 86.5% | 83.3% to 89.2% |
| *>13 years* | 124/188 | 66.3% | 59.1% to 73.0% | 70.2% | 62.0% to 77.5% |
| *13-17 years* | 179/230 | 77.8% | 71.9% to 83.0% | 82.8% | 75.8% to 88.8% |
| *18-50 years* | 2261/2750 | 82.2% | 80.7% to 83.6% | 87.5% | 84.1% to 90.5% |
| *>50 years* | 847/1043 | 81.2% | 78.7% to 83.5% | 86.4% | 82.3% to 89.9% |
| *Blantyre* | 678/768 | 88.3% | 85.8% to 90.5% | 93.9% | 89.7% to 97.5% |
| *Dedza* | 406/527 | 77.0% | 73.2% to 80.6% | 81.9% | 76.9% to 86.3% |
| *Dowa* | 334/441 | 75.7% | 75.7% to 79.7% | 80.6% | 75.1% to 85.2% |
| *Kasungu* | 486/645 | 75.3% | 71.8% to 78.6% | 80.2% | 75.4% to 84.3% |
| *Lilongwe* | 1132/1327 | 85.3% | 83.3% to 87.2% | 90.8% | 86.9% to 94.0% |
| *Machinga* | 380/477 | 79.7% | 75.8% to 83.2% | 84.8% | 79.6% to 89.2% |
| *Zomba* | 338/434 | 77.9% | 73.7% to 81.7% | 82.9% | 77.4% to 87.5% |
| *Urban* | 1252/1373 | 91.2% | 89.6% to 92.6% | 97.0% | 93.6% to 99.6% |
| *Rural* | 896/1156 | 77.5% | 75.0% to 79.9% | 82.5% | 78.5% to 85.9% |

\* The sensitivity and specificity of the assay as independently validated are 94% [95% *CI* 0.91 to 0.97]) and 100% [95% *CI* 0.99 to 1.00]), respectively

Participants residing in urban areas were more likely to test seropositive than those from rural areas (91% vs. 78%; adjusted odds ratio, 2.81; 95% CI, 2.20 to 3.62) (**Table 3**). Males were less likely to be seropositive than females (78% vs 84%; adjusted odds ratio, 0.61; 95% CI, 0.52 to 0.72). The seroprevalence among the age groups did not vary greatly among those aged 13 years and above; but it was lowest in children younger than 13 years of age (66%) and highest among adults 18 to 50 years of age (82%). Children 13 to 17 years of age were more likely to be seropositive than children younger than 13 years of age (78% vs. 66%; adjusted odds ratio, 1.80; 95% CI, 1.15 to 2.83). Participants who had received a COVID-19 vaccine were more likely to be seropositive than unvaccinated participants (1 dose vs no dose, 94% vs. 77%; adjusted odds ratio, 4.83; 95% CI, 3.40 to 7.10; 2 doses vs no dose, 97% vs. 77%; adjusted odds ratio, 6.65; 95% CI, 4.16 to 11.4).

**Table 3.**
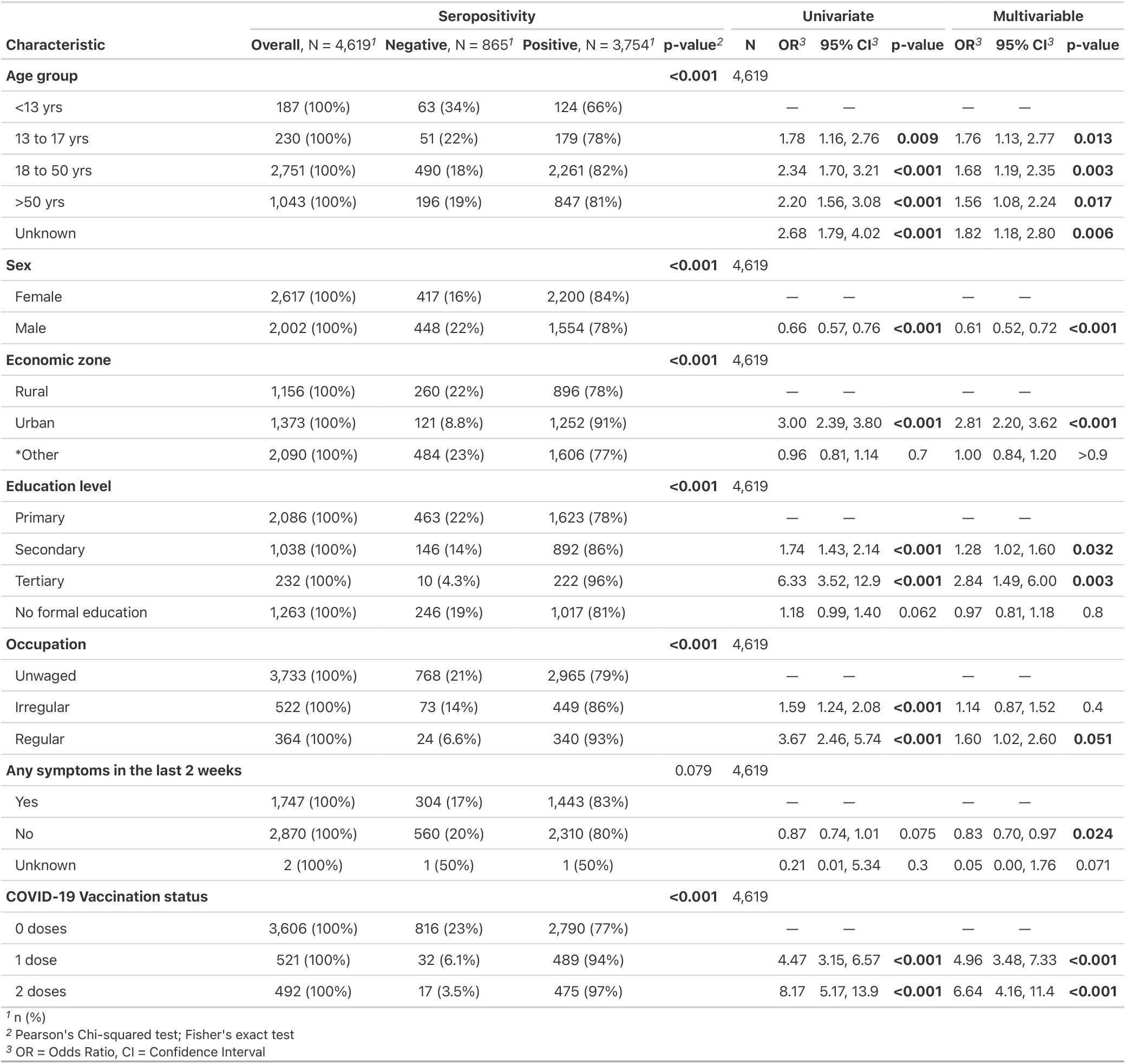
Factors associated with SARS-CoV-2 antibody seropositivity.

| Characteristic | Seropositivity |  |  |  | Univariate |  |  |  | Multivariable |  |  |
| --- | --- | --- | --- | --- | --- | --- | --- | --- | --- | --- | --- |
|  | Overall, N = 4,619<sup>1</sup> | Negative, N = 865<sup>1</sup> | Positive, N = 3,754<sup>1</sup> | p-value<sup>2</sup> | N | OR<sup>3</sup> | 95% CI<sup>3</sup> | p-value | OR<sup>3</sup> | 95% CI<sup>3</sup> | p-value |
| **Age group** |  |  |  | **<0.001** | 4,619 |  |  |  |  |  |  |
| <13 yrs | 187 (100%) | 63 (34%) | 124 (66%) |  |  | — | — |  | — | — |  |
| 13 to 17 yrs | 230 (100%) | 51 (22%) | 179 (78%) |  |  | 1.78 | 1.16, 2.76 | **0.009** | 1.76 | 1.13, 2.77 | **0.013** |
| 18 to 50 yrs | 2,751 (100%) | 490 (18%) | 2,261 (82%) |  |  | 2.34 | 1.70, 3.21 | **<0.001** | 1.68 | 1.19, 2.35 | **0.003** |
| >50 yrs | 1,043 (100%) | 196 (19%) | 847 (81%) |  |  | 2.20 | 1.56, 3.08 | **<0.001** | 1.56 | 1.08, 2.24 | **0.017** |
| Unknown |  |  |  |  |  | 2.68 | 1.79, 4.02 | **<0.001** | 1.82 | 1.18, 2.80 | **0.006** |
| **Sex** |  |  |  | **<0.001** | 4,619 |  |  |  |  |  |  |
| Female | 2,617 (100%) | 417 (16%) | 2,200 (84%) |  |  | — | — |  | — | — |  |
| Male | 2,002 (100%) | 448 (22%) | 1,554 (78%) |  |  | 0.66 | 0.57, 0.76 | **<0.001** | 0.61 | 0.52, 0.72 | **<0.001** |
| **Economic zone** |  |  |  | **<0.001** | 4,619 |  |  |  |  |  |  |
| Rural | 1,156 (100%) | 260 (22%) | 896 (78%) |  |  | — | — |  | — | — |  |
| Urban | 1,373 (100%) | 121 (8.8%) | 1,252 (91%) |  |  | 3.00 | 2.39, 3.80 | **<0.001** | 2.81 | 2.20, 3.62 | **<0.001** |
| *Other | 2,090 (100%) | 484 (23%) | 1,606 (77%) |  |  | 0.96 | 0.81, 1.14 | 0.7 | 1.00 | 0.84, 1.20 | >0.9 |
| **Education level** |  |  |  | **<0.001** | 4,619 |  |  |  |  |  |  |
| Primary | 2,086 (100%) | 463 (22%) | 1,623 (78%) |  |  | — | — |  | — | — |  |
| Secondary | 1,038 (100%) | 146 (14%) | 892 (86%) |  |  | 1.74 | 1.43, 2.14 | **<0.001** | 1.28 | 1.02, 1.60 | **0.032** |
| Tertiary | 232 (100%) | 10 (4.3%) | 222 (96%) |  |  | 6.33 | 3.52, 12.9 | **<0.001** | 2.84 | 1.49, 6.00 | **0.003** |
| No formal education | 1,263 (100%) | 246 (19%) | 1,017 (81%) |  |  | 1.18 | 0.99, 1.40 | 0.062 | 0.97 | 0.81, 1.18 | 0.8 |
| **Occupation** |  |  |  | **<0.001** | 4,619 |  |  |  |  |  |  |
| Unwaged | 3,733 (100%) | 768 (21%) | 2,965 (79%) |  |  | — | — |  | — | — |  |
| Irregular | 522 (100%) | 73 (14%) | 449 (86%) |  |  | 1.59 | 1.24, 2.08 | **<0.001** | 1.14 | 0.87, 1.52 | 0.4 |
| Regular | 364 (100%) | 24 (6.6%) | 340 (93%) |  |  | 3.67 | 2.46, 5.74 | **<0.001** | 1.60 | 1.02, 2.60 | **0.051** |
| **Any symptoms in the last 2 weeks** |  |  |  | 0.079 | 4,619 |  |  |  |  |  |  |
| Yes | 1,747 (100%) | 304 (17%) | 1,443 (83%) |  |  | — | — |  | — | — |  |
| No | 2,870 (100%) | 560 (20%) | 2,310 (80%) |  |  | 0.87 | 0.74, 1.01 | 0.075 | 0.83 | 0.70, 0.97 | **0.024** |
| Unknown | 2 (100%) | 1 (50%) | 1 (50%) |  |  | 0.21 | 0.01, 5.34 | 0.3 | 0.05 | 0.00, 1.76 | 0.071 |
| **COVID-19 Vaccination status** |  |  |  | **<0.001** | 4,619 |  |  |  |  |  |  |
| 0 doses | 3,606 (100%) | 816 (23%) | 2,790 (77%) |  |  | — | — |  | — | — |  |
| 1 dose | 521 (100%) | 32 (6.1%) | 489 (94%) |  |  | 4.47 | 3.15, 6.57 | **<0.001** | 4.96 | 3.48, 7.33 | **<0.001** |
| 2 doses | 492 (100%) | 17 (3.5%) | 475 (97%) |  |  | 8.17 | 5.17, 13.9 | **<0.001** | 6.64 | 4.16, 11.4 | **<0.001** |

<sup>1</sup> n (%)
<sup>2</sup> Pearson's Chi-squared test; Fisher's exact test
<sup>3</sup> OR = Odds Ratio, CI = Confidence Interval

Participants who reported COVID-19 symptoms in the last two weeks, including persistent cough, loss of taste, sore throat, chest pain, myalgia, shortness of breath, severe/unusual headache, fatigue, fever and runny/blocked nose, were more likely to be seropositive than participants who did not report any symptoms (83% vs. 80%; adjusted odds ratio, 1.21; 95% CI, 1.03 to 1.42) (**Table 3**). Participants who reached secondary (86 % vs. 78 %; adjusted odds ratio, 1.27; 95% CI, 1.02 to 1.59) and tertiary (96 % vs. 78 %; adjusted odds ratio, 2.83; 95% CI, 1.49 to 5.99) education levels had a higher seroprevalence than participants at primary education level. Participants with a regular waged job were more likely to be seropositive than unwaged participants (93 % vs. 79 %; adjusted odds ratio, 1.60; 95% CI, 1.02 to 2.61).

### COVID-19 vaccination

Of the 4,619 participants who were included in the data analysis, 3,794 of them had known age and were eligible for COVID-19 vaccination at the time of the study being 18 years and older (**Table 4**). Overall, 23% (878/3,794) of the participants had received at least one dose of the COVID-19 vaccine, with 12% (458/3,794) receiving 1 dose and 11% (420/3,794) received 2 doses. The age group of 50 years and above had a higher proportion of vaccinated individuals than those between 18 to 49 years (33% vs. 19%, p<0.001). Moreover, the age group of 50 years and above had a higher proportion of vaccinated individuals that received 2 doses than those between the ages of 18 to 49 years (19% vs. 8%).

**Table 4.** COVID-19 vaccination.

| Characteristic | Overall, N = 3,794<sup>1</sup> | 0 doses, N = 2,916<sup>1</sup> | 1 dose, N = 458<sup>1</sup> | 2 doses, N = 420<sup>1</sup> | p-value<sup>2</sup> |
| --- | --- | --- | --- | --- | --- |
| **Age group** |  |  |  |  | **<0.001** |
| 18 to 50 yrs | 2,751 (100%) | 2,216 (81%) | 312 (11%) | 223 (8.1%) |  |
| >50 yrs | 1,043 (100%) | 700 (67%) | 146 (14%) | 197 (19%) |  |
| **Sex** |  |  |  |  | **0.011** |
| Female | 2,176 (100%) | 1,689 (78%) | 274 (13%) | 213 (9.8%) |  |
| Male | 1,618 (100%) | 1,227 (76%) | 184 (11%) | 207 (13%) |  |
| **HIV status** |  |  |  |  | **0.6** |
| HIV- | 3,668 (100%) | 2,824 (77%) | 441 (12%) | 403 (11%) |  |
| HIV+ | 126 (100%) | 92 (73%) | 17 (13%) | 17 (13%) |  |
| **Location** |  |  |  |  | **<0.001** |
| Blantyre | 611 (100%) | 476 (78%) | 44 (7.2%) | 91 (15%) |  |
| Dedza | 468 (100%) | 394 (84%) | 61 (13%) | 13 (2.8%) |  |
| Dowa | 308 (100%) | 208 (68%) | 83 (27%) | 17 (5.5%) |  |
| Kasungu | 538 (100%) | 382 (71%) | 105 (20%) | 51 (9.5%) |  |
| Lilongwe | 1,124 (100%) | 834 (74%) | 109 (9.7%) | 181 (16%) |  |
| Machinga | 401 (100%) | 364 (91%) | 14 (3.5%) | 23 (5.7%) |  |
| Zomba | 344 (100%) | 258 (75%) | 42 (12%) | 44 (13%) |  |
| **Education level** |  |  |  |  | **<0.001** |
| Primary | 1,602 (100%) | 1,265 (79%) | 202 (13%) | 135 (8.4%) |  |
| Secondary | 910 (100%) | 685 (75%) | 95 (10%) | 130 (14%) |  |
| Tertiary | 206 (100%) | 128 (62%) | 20 (9.7%) | 58 (28%) |  |
| No formal education | 1,076 (100%) | 838 (78%) | 141 (13%) | 97 (9.0%) |  |
| **Occupation** |  |  |  |  | **<0.001** |
| Irregular | 462 (100%) | 368 (80%) | 45 (9.7%) | 49 (11%) |  |
| Regular | 308 (100%) | 169 (55%) | 31 (10%) | 108 (35%) |  |
| Unwaged | 3,024 (100%) | 2,379 (79%) | 382 (13%) | 263 (8.7%) |  |
| **Economic zone** |  |  |  |  | **<0.001** |
| Urban | 1,123 (100%) | 855 (76%) | 82 (7.3%) | 186 (17%) |  |
| Rural | 956 (100%) | 713 (75%) | 113 (12%) | 130 (14%) |  |
| *Other | 1,715 (100%) | 1,348 (79%) | 263 (15%) | 104 (6.1%) |  |

<sup>1</sup> n (%)
<sup>2</sup> Pearson's Chi-squared test
\* includes other districts apart from city-harboring districts of Lilongwe, Blantyre and Zomba

All major cities of Lilongwe (16%), Blantyre (15%) and Zomba (13%), had a greater proportion of individuals that received two doses of COVID-19 vaccine compared to the other districts (all < 9.6%). Moreover, the proportion of 2-dose vaccinated individuals was higher in urban areas than rural areas (17% vs 14%, p<0.001). A greater proportion of participants who had gone up to tertiary education level (28%) received two doses of the COVID-19 vaccine, whereas those at primary and secondary level were 8% and 14%, respectively (p<0.001). The proportion of 2-dose vaccinated people were higher (p<0.001) among those with a regular wage (35%) compared to unwaged (9%) or those with an irregular wage (11%).

### National COVID-19 epidemiological trends

Our study was conducted during the fourth wave, which was dominated by the Omicron variant ^18,19^. During this wave, the daily case incidence increased more rapidly and also decreased more quickly compared to the third (Delta) wave, with the peak in July 2021 and December 2021, respectively (**Figure 2**). The number of confirmed clinical cases at the peak of the third wave were 13300 and during the fourth wave were 14298, whereas the peak episodes of hospitalization were 1429 and 695, while deaths were 463 and 164, respectively. The proportion of hospitalisation within the confirmed clinical cases was two-fold lower in the fourth wave compared to the third wave (4.86% (95% CI, 4.52 to 5.23) (695/14298) vs 10.7% (95% CI, 10.2 to 11.3) (1429/13330), p<0.0001). While, the proportion of deaths within the confirmed clinical cases was three-fold lower in the fourth wave compared to the third wave (1.15% (95% CI, 1.00 to 1.34) (164/14298) vs 3.48% (95% CI, 3.18 to 3.81) (463/13330), p<0.0001).

**Figure 2.**
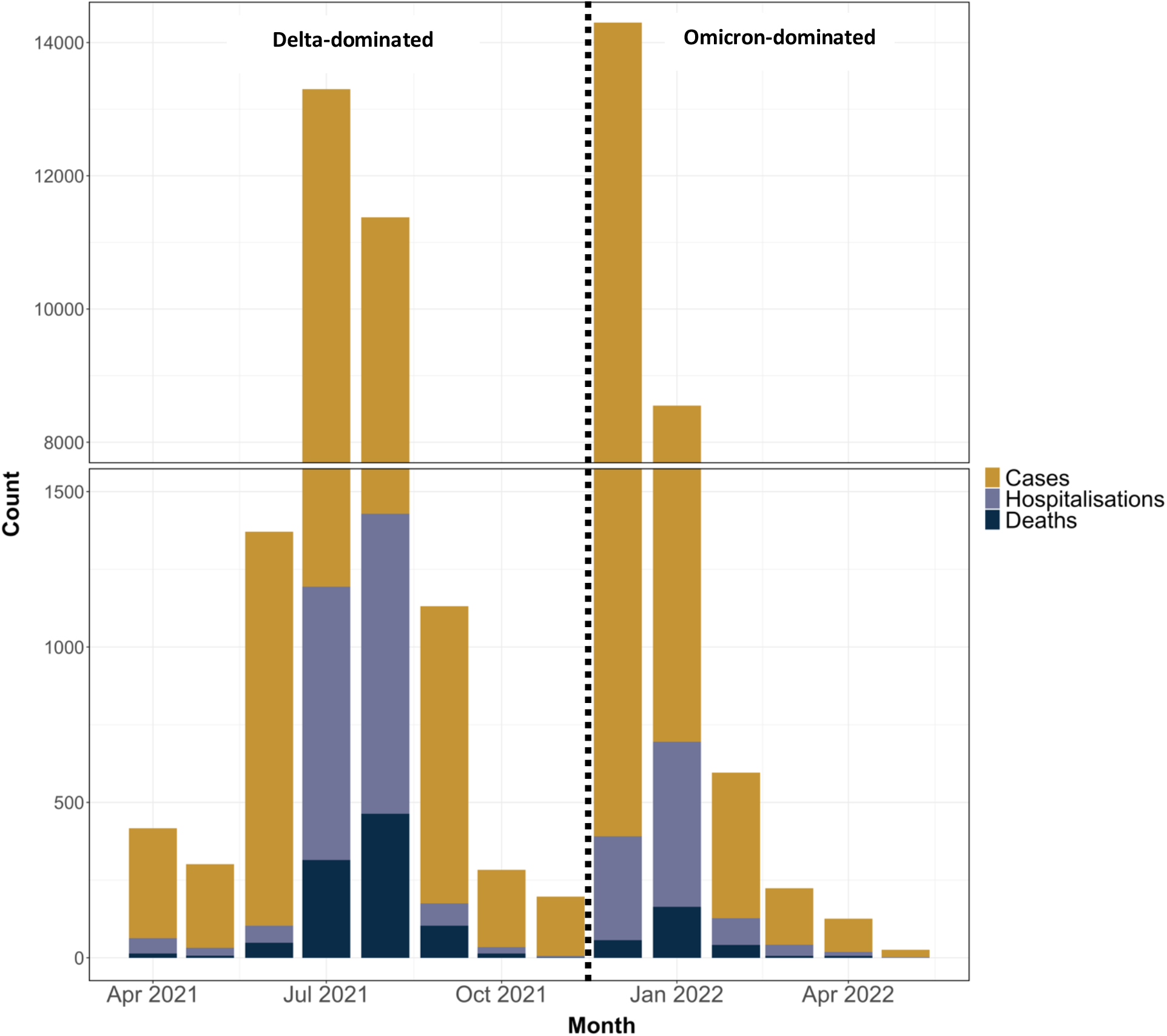
COVID-19 Cases, Hospitalizations, and Deaths in Malawi, from April 2021 through April 2022. Shown are incidences of monthly cases, hospitalizations, and deaths attributable to coronavirus disease 2019 (COVID-19). The data were sourced from the Public Health Institute of Malawi (PHIM). Cases included asymptomatic and symptomatic infections with severe acute respiratory syndrome coronavirus 2 (SARS-CoV-2) confirmed by either a nucleic acid amplification assay or a rapid antigen test. Hospitalizations included admissions for SARS-CoV-2 infection, as well as admissions for other illnesses in which SARS-CoV-2 infection was incidentally identified on routine screening at the time of admission. Death attributable to COVID-19 was defined as a death resulting from a clinically compatible illness in a probable or confirmed COVID-19 case unless there is a clear alternative cause of death that cannot be related to COVID disease (e.g., trauma), according to the national guidelines

## Discussion

The results of our study showed high SARS-CoV-2 seropositivity of 86% across the population during the Omicron B.1.1.529 variant-dominated fourth wave. This high seroprevalence was primarily due to prior SARS-CoV-2 infection, as evidence by the low vaccine uptake in this population and the 70% seroprevalence among participants who had not received a COVID-19 vaccine. Against this background, we observed reduction in the proportion of hospitalization and deaths from the reported infections during the Omicron-dominated fourth wave, compared to the Delta-dominated third wave. This is in support of Madhi *et al* that have observed a similar phenomenon in South Africa, which they termed “decoupling” ^20^.

Notwithstanding the reduced number of hospitalization and deaths in Malawi, the Omicron variant drove a rapid rise in SARS-CoV-2 infections consistent with its high transmissibility ^21,22^ as observed in other settings ^1,23^. Based on previous estimates from our adult blood donor serosurvey, the estimated seroprevalence by July 2021 was >65% ^10^, reaching around 80% by November 2021 (unpublished), before the Omicron-dominant fourth wave in Malawi. In South Africa, a lower rate of hospitalization and deaths was associated with the high seroprevalence before the onset of the Omicron-dominant wave, driven by both vaccination and infection ^1^. In Malawi, there was at least a two-fold reduction in proportion of COVID-19 hospitalizations and deaths among the clinical cases reported in the fourth compared to third wave. Madhi *et al*. suggests that, in South Africa, cell-mediated immunity induced through prior infection and vaccination contributed to protective immunity against severe disease ^1^. This suggestion was based on earlier evidence that showed that the Omicron variant highly evades neutralization antibody activity ^5-7,24^ but is ably cross-recognized by T cells ^25,26^. Evidence of T cell-mediated protection against COVID-19 is accumulating and is consistent with their role in viral clearance, where they might not directly prevent infection but halt progression of infection to disease ^27,28^.

The SARS-CoV-2 seroprevalence was generally high across all the seven districts surveyed in the current study, with higher seroprevalence in urban compared to rural areas, as reported in our blood donor serosurvey ^10^ and in other sub-Saharan African settings ^29^. This is consistent with national COVID-19 data at the time of the study that reported 51% of confirmed cases in the country were from the cities of Blantyre and Lilongwe ^9^, with a caveat that rural areas had relatively fewer SARS-CoV-2 testing sites. The high seroprevalence seen in the urban areas could be attributed to these areas being hotspots for SARS-CoV-2 transmission. Individuals residing in the urban areas were more likely to have attained higher level education and have regular waged jobs, likely increasing their risk of exposure due to increased interaction with others, closed workspaces, frequent travel on public transport and international travel that also increases the risk of exposure to new variants. Supporting this, seroprevalence was higher in those with secondary or tertiary level education, and in those on regular waged jobs. Our study also found men to have a lower seropositivity, which is contrary to male-dominated reported national COVID-19 cases, this could suggest that females experienced less severe forms of COVID-19 to trigger testing or were less likely to present for testing. These findings highlight the need to tailor public health measures according to context as the pandemic progresses.

Our study has some limitations. First, we used an anti-RBD ELISA assay that was developed against the wild type SARS-CoV-2, hence the extensive mutations in the RBD in the Omicron variant could result in false negative results in individuals who were previously or actively infected with this variant.

Nevertheless, considering the extremely high seroprevalence, our conclusions are unlikely to be significantly impacted by the theoretical possibility of lower assay specificity. Second, the use of publicly available reported national COVID-19 surveillance data has its own caveats, including under ascertainment of the cases and deaths or changes in testing strategies over time, and this needs to be taken into consideration when interpreting the comparisons across the waves. However, to our knowledge, there were no significant changes in the clinical management policy of severe COVID-19 cases between the third and fourth waves, hence the comparison between the waves is likely accurate.

Thirdly, we had a relatively small numbers of children to give precise estimates of seroprevalence in this population. Finally, the study was conducted and concluded before the end of the fourth wave, hence this could result in seroprevalence estimates being an underestimate of the actual cumulative exposure for Malawi.

The reduction in the proportion of hospitalizations and deaths from COVID-19 during the Omicron-dominant fourth wave in Malawi, in the context of low vaccination coverage and high infection-induced seroprevalence, could be a watershed moment for the COVID-19 pandemic in under-vaccinated countries like Malawi. However, the number of COVID-19 preventable deaths are still significant and a greater focus on identifying and protecting those most at risk is increasingly important. Up until a sarbecoviruses or pan-coronavirus vaccine is available, to maximise protection against severe disease in high seroprevalence and resource-constrained settings like Malawi, there might be the need to amend vaccination policy of current COVID-19 vaccines from mass vaccination campaigns to targeted vaccination that focuses on at-risk populations including the elderly and those with co-morbidities.

## Data Availability

All data produced in the present study are available upon reasonable request to the authors

## Acknowledgements

The authors thank all participants whose samples are used in this study. We thank all the district teams and leadership for their support and contribution to this study. We also thank Mtisunge Yelewa, Selemani Ngwira, Eranive Chipungu, Triza Chirwa, and Sikhona Chipeta. We acknowledge use of the WHO Unity protocols for this study.

## Author contributions

Conceptualization of study: ACM, WK, OD, DK, JP, CM

Study design: ACM, WK, OD, DK, JP, CM, MK, ND

Data collection: ULM, DM, KN

Laboratory testing: JM, MM, IC, BO

Data analysis: KCJ1, ULM, JM

Accessed and verified the underlying data: KCJ1, MK

Study management: ULM, KN

Initial manuscript draft: ULM, KCJ1, ACM

Final manuscript writing: ULM, KCJ1, ACM, RSH, TDS, KCJ2, NF, ASA, AC

Manuscript review and approval: All authors

